# Separating Clinical and Subclinical Depression by Big Data Informed Structural Vulnerability Index and Its impact on Cognition: ENIGMA Dot Product

**DOI:** 10.1101/2021.09.18.21263763

**Authors:** Peter Kochunov, Yizhou Ma, Kathryn S. Hatch, Lianne Schmaal, Neda Jahanshad, Paul M. Thompson, Bhim M. Adhikari, Heather Bruce, Joshua Chiappelli, Andrew Van der vaart, Eric L. Goldwaser, Aris Sotiras, Tianzhou Ma, Shuo Chen, Thomas E. Nichols, L. Elliot Hong

## Abstract

Big Data neuroimaging collaborations including Enhancing Neuro Imaging Genetics through Meta-Analysis (ENIGMA) integrated worldwide data to identify regional brain deficits in major depressive disorder (MDD). We evaluated the sensitivity of translating ENIGMA-defined MDD deficit patterns to the individual level. We treated ENIGMA MDD deficit patterns as a vector to gauge the similarity between individual and MDD patterns by calculating ENIGMA dot product (EDP). We analyzed the sensitivity and specificity of EDP in separating subjects with (1) subclinical depressive symptoms without a diagnosis of MDD, (2) single episode MDD, (3) recurrent MDD, and (4) controls free of neuropsychiatric disorders. We compared EDP to the Quantile Regression Index (QRI; a linear alternative to the brain age metric) and the global gray matter thickness and subcortical volumes and fractional anisotropy (FA) of water diffusion. We performed this analysis in a large epidemiological sample of UK Biobank (UKBB) participants (N=17,053/19,265 M/F). Group-average increases in depressive symptoms from controls to recurrent MDD was mirrored by EDP (r2=0.85), followed by FA (r2=0.81) and QRI (r2=0.56). Subjects with MDD showed worse performance on cognitive tests than controls with deficits observed for 3 out of 9 cognitive tests administered by the UKBB. We calculated correlations of EDP and other brain indices with measures of cognitive performance in controls. The correlation pattern between EDP and cognition in controls was similar (r2=0.75) to the pattern of cognitive differences in MDD. This suggests that the elevation in EDP, even in controls, is associated with cognitive performance - specifically in the MDD-affected domains. That specificity was missing for QRI, FA or other brain imaging indices. In summary, translating anatomically informed meta-analytic indices of similarity using a linear vector approach led to better sensitivity to depressive symptoms and cognitive patterns than whole-brain imaging measurements or an index of accelerated aging.

## 1. Introduction

Major depressive disorder (MDD) is the most common of all severe mental illnesses affecting up to 20-30% of the worldwide population and inflicting a tremendous burden on patients, caregivers, and communities. The onset and course of MDD are influenced by genetic and environmental risk factors that are complex and remain poorly understood. Moreover, clinical and neuroimaging findings in MDD have historically been affected by heterogeneity and poor reproducibility (1). Collaborative studies by the Enhancing Neuro Imaging Genetics Meta Analyses (ENIGMA) led to a neuroimaging initiative to overcome the historically high heterogeneity and lack of consistent findings in MDD (2-8). The standardized workflows developed by ENIGMA made it possible to elucidate the imaging patterns associated with MDD in cohorts across the world. The inclusive worldwide nature of the collaboration has reduced site-specific variances and biases in diagnosis, inclusion and exclusion criteria, demographics medication, and environment. These concerted efforts yielded the discovery of deficit patterns that are shared by patients worldwide and remain present even after treatment with existing therapies.

In schizophrenia research, ENIGMA efforts led to the development of the similarity indices that use ENIGMA deficit patterns as the basis for vulnerability measurements (9). It is unknown, however, to what extent this approach can also be applied to MDD – specifically at different levels of severity including recurrent, single-episode, and subthreshold illness. It is also unclear if the better alignment with disordered patterns can explain variance in the cognition as cognitive deficits in MDD contribute to the burden of this disorder. Finally, it remains unclear if this approach distinguishes MDD groups better than multivariate metrics based on deviations from normative reference distributions (e.g., ‘brain age’ metrics) (10, 11). We design this study to answer two questions: A) is the similarity to the characteristic patterns of regional deficits derived by ENIGMA-MDD a better predictor of clinical features of depression, as compared to averaged brain structural measures? B) can the similarity index explain the cognitive variance in patients and non-psychiatric controls? We evaluated a linear algebra approach to calculate Enigma Dot Product (EDP) between the vector of individual brain structural measurements and the ENIGMA-derived patterns for MDD in the high-dimensional space of brain structural measurements. A larger EDP value suggests that the individual brain structural vector deviates from the expected population mean in the direction defined by the ENIGMA MDD deficit vector.

The neuroimaging structures used for the EDP calculation included 33 cortical thickness measures, 24 white matter diffusion tensor imaging (DTI) measures, and 7 subcortical gray matter volume measures. To compare EDP with traditional structural measures, we selected the whole-brain average measures including whole-brain average gray matter (GM) cortical thickness, subcortical GM volumes and fractional anisotropy (FA) of water diffusion. We also selected a Quantile Regression Index (QRI) - a linear analog of the BrainAge index that was shown to be sensitive to accelerated brain aging trends in MDD (12). We performed these analyses in the UK Biobank cohort which was chosen because it is the largest publicly available dataset with high-quality brain MRI scans. Notably, the UKBB participants were not included in the ENIGMA MDD studies and therefore it is an independent source of test data for the questions tackled here. We classified the UKBB participants into four subgroups that reflect severity of depression. These included (A) people who express some subclinical depression symptoms without a lifetime history of diagnosed MDD at the time of scan (B) people who had a single probable depressive episode, and (C) those who suffered from more than one episode of MDD (termed recurrent MDD). As a fourth comparison group, we also carefully chose a group of healthy controls (HC) who were free of any neurological or psychiatric disorders and denied significant depressive symptoms in their lifetime.

We hypothesized that alignment with expected MDD pattern would rise from controls to subclinical, single, and recurrent MDD. We tested if the group differences in EDP, the whole-brain averaged structural measures and accelerated aging index would provide the best predictions that are proportional to the group differences in symptoms. We further hypothesized that the EDP would predict subclinical cognitive deficits in individuals without a formal diagnosis of MDD better than the whole brain averaged structural measures. We examined a novel proposition that EDP can predict cognitive deficits in non-psychiatric controls, and specifically in domains affected by MDD, and would do so better than the whole brain averaged structural measures. The overall goal of our study was to examine to what extent a linear combinatoric index of structural measures could be used to complement traditional structural measures and to provide an even more powerful approach to better understand the clinical differentiation and neurobiological underpinnings of different levels of depression.

## 2. Methods

### 2.1 Participants

Neuroimaging and clinical data, including structural and diffusion imaging phenotypes, were available for N=37,285 individuals (17,082/20,203 M/F). Data were collected between 2012 and 2019 in participants recruited in the United Kingdom (13). All participants provided written informed consent.

### 2.2 Major Depressive Disorder Classification

We classified participants into four subgroups of depression, i.e., subclinical depression without a current or lifetime MDD diagnosis (subclinical depression N=8,822; 4,313/4,509 M/F), single episode (N=1,147; 443/704 M/F), and recurrent MDD (N=8,089; 2,971/ 5,118 M/F) (Table 1). We integrated classification criteria previously published for participants’ responses to the touchscreen questionnaire at each visit (Smith et al., 2013) with hospital records and self-reported diagnoses. Details of the classification are listed in Table 2. We took a conservative approach to defining controls so that participants with missing values (and thus preventing a definitive conclusion on certain criterion) were not used. We also excluded participants with significant neurological and psychiatric conditions (including bipolar disorder) and brain cancer per hospital records, self-report and probable bipolar disorder per (14).

**Table 1.** Classification of participants into subgroups of depression and non-psychiatric controls.

| MDD Classification | Number of Participants (M/F) | Average Age ± SD |
| --- | --- | --- |
| Subclinical | 8,822 (4,313/4,509) | $63.6 \pm 7.5$ |
| Single Episode | 1,147 (443/704) | $63.8 \pm 7.4$ |
| Recurrent | 8,089 (2,971/ 5,118) | $62.1 \pm 7.3$ |
| Controls | 7,266 (4,033/3,233) | $65.1 \pm 7.5$ |
| Total | 37285 (17082/20203) | $63.6 \pm 7.5$ |

We used the UKBB parser software (https://github.com/USC-IGC/ukbb_parser) to collect a group of N=7,266 (4,033/3,233 M/F, age = 63.3±7.5) in the overall sample who were free of ICD codes corresponding to any neurological or psychiatric illnesses and major health conditions. The exclusion criteria included: bipolar disorder, schizophrenia, anxiety, Alzheimer’s disease, head trauma, stroke, Parkinson’s, PTSD, meningitis, multiple sclerosis, migraines, and other demyelinating diseases, cancer, heart disease, and were treated as non-neuropsychiatric controls (Table 1).

### 2.3 Imaging Protocol and Processing

In this study, we examined regional cortical gray matter thickness, subcortical gray matter structural volume and tract-wise measures of fractional anisotropy (FA) values provided by the UKBB. These phenotypes were extracted from neuroimaging data collected with Siemens Skyra 3T scanners using standard 32-channel radiofrequency (RF) head coils. The brain imaging protocol collected high resolution T1-weighted 3D MP-RAGE scans (resolution=1×1×1 mm, FOV=208×256×256, duration=5 minutes, sagittal, in-plane acceleration iPAT=2, prescan-normalize). Diffusion data were collected with a resolution of 2×2×2 mm and two diffusion shells of *b*=1000 and 2000 s/mm^2^ with 50 diffusion directions per shell and 5 *b*=0 images (FOV=104×104×72, duration=7 minutes).

EDP calculations were based on the meta-analytical pattern aggregated from independent imaging studies of an illness to establish the ‘gold standard’ findings for that illness – here, the corresponding imaging measurements from ENIGMA were used. Accordingly, all imaging data were processed using the UKBB workflow that is based on the ENIGMA imaging processing pipelines. Details of the image preprocessing and analysis are provided by UKBB (biobank.ctsu.ox.ac.uk/crystal/crystal/docs/brain_mri.pdf). Briefly, the UKBB workflow provides brain imaging measurements that have been evaluated across many ENIGMA studies. These phenotypes included 24 regional white matter tract FA values, 33 regional estimates of cortical gray matter (GM) thickness, volumes of the lateral ventricles, and 7 subcortical gray matter volumes per hemisphere that corresponded to those derived by ENIGMA workflows. Measures from the left and right structures were averaged.

### 2.4 Calculation of linear indices of similarity

All linear indices of vulnerability were calculated using the ‘RVIpkg’ implemented in [R] software in Kochunov et al. (2020). Briefly, the effects of age, sex, and the intracranial volume were regressed. The inverse-normal transformation was applied to the residuals. This produced a vector of Z-values for everyone, the absolute value of which was the deviation of the individual from the mean of the group. The EDP was calculated as the normalized dot product between P and E using the following equation.

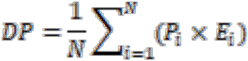

Where P_i_ was the vector of deviation from the mean for the *i*-th phenotype and E_i_ was the vector of meta-analytical effect size for the *i*-th phenotype for MDD provided by ENIGMA. N was the total number of imaging phenotypes.

### 2.5 Calculation of QRI

Individual assessments of brain aging were performed using quantile regression analysis to derive a quantile regression index (QRI) score provided in the ‘QRIpkg’ for R (15) available for download at https://CRAN.R-project.org/package=QRIpkg. Briefly, the normative modeling technique provided a map of *individual* deviations from the expected aging trends using the whole sample. The QRI function used a quantile regression analysis with age serving as a predictor for cortical regional gray matter thickness, subcortical gray matter structure volumes, and white matter fractional anisotropy values to fit three separate models for the 5^th^, 50^th^, and 95^th^ percentiles. Then, values for each individual subject were compared to the expected aging trajectory, derived from the whole sample, and each regional measure was assigned a score: values >95% of the expected age data were assigned a value of “-1”, indicating an individual’s actual brain age was significantly younger than what was expected for that age; values <5% received a value of “1”, indicating an individual’s actual brain age was significantly older than what was expected for that age; all others were assigned “0”. Regional scores were averaged for cortical thickness, subcortical volume, and white matter to create tissue specific QRIs. A whole-brain QRI was derived by averaging the three-tissue specific QRIs.

### 2.7 Cognitive assessment

Participants completed the automated UK Biobank cognitive test battery on a touchscreen computer (16). We included all tests collected by the UKBB covering cognitive domains of processing speed, cognitive flexibility, working memory, visuospatial learning/memory, perceptual reasoning, executive functioning/planning, and fluid intelligence.

### 2.8 Statistics

All statistical analyses were performed using RStudio v3.6.3 [71]. All measurements were preprocessed by correcting for age and sex prior to analysis. The relative effects of DP and other measurements on cognition were tested using the following model:

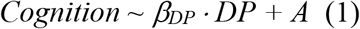

Where *β* was a linear predictor coefficient, and A was the intercept. Model 1 was used in controls to examine whether being merely similar to MDD in brain patterns was associated with poorer cognitive performance and whether the pattern of this association matched the cognitive differences between subjects with MDD and controls.

## 3. Results

### 3.1 Group differences in symptoms and biomarkers

The four subject groups demonstrated the expected difference in symptom severity (Figure 1). The ENIGMA dot product (EDP) showed the average values for four groups that were significantly higher in the clinical and subclinical depression groups than in controls (Figure 1). The average EDP values per group were correlated with the groups’ average symptom severity (r^2^=0.85). In comparison, among the three whole-brain average indices, only the average FA value showed a pattern that agreed with the average symptoms for each group (r^2^=0.81). Individual depressive symptoms in subjects with depression showed a significant correlation for EDP (r=0.04, t=6.4, p=10^−10^), followed by average FA (r=0.03, t=4.7, p=10^−6^) and QRI (r=0.02, t=4.2, p=10^−5^) and but no significant correlation for GM thickness and volume.

**Figure 1.**
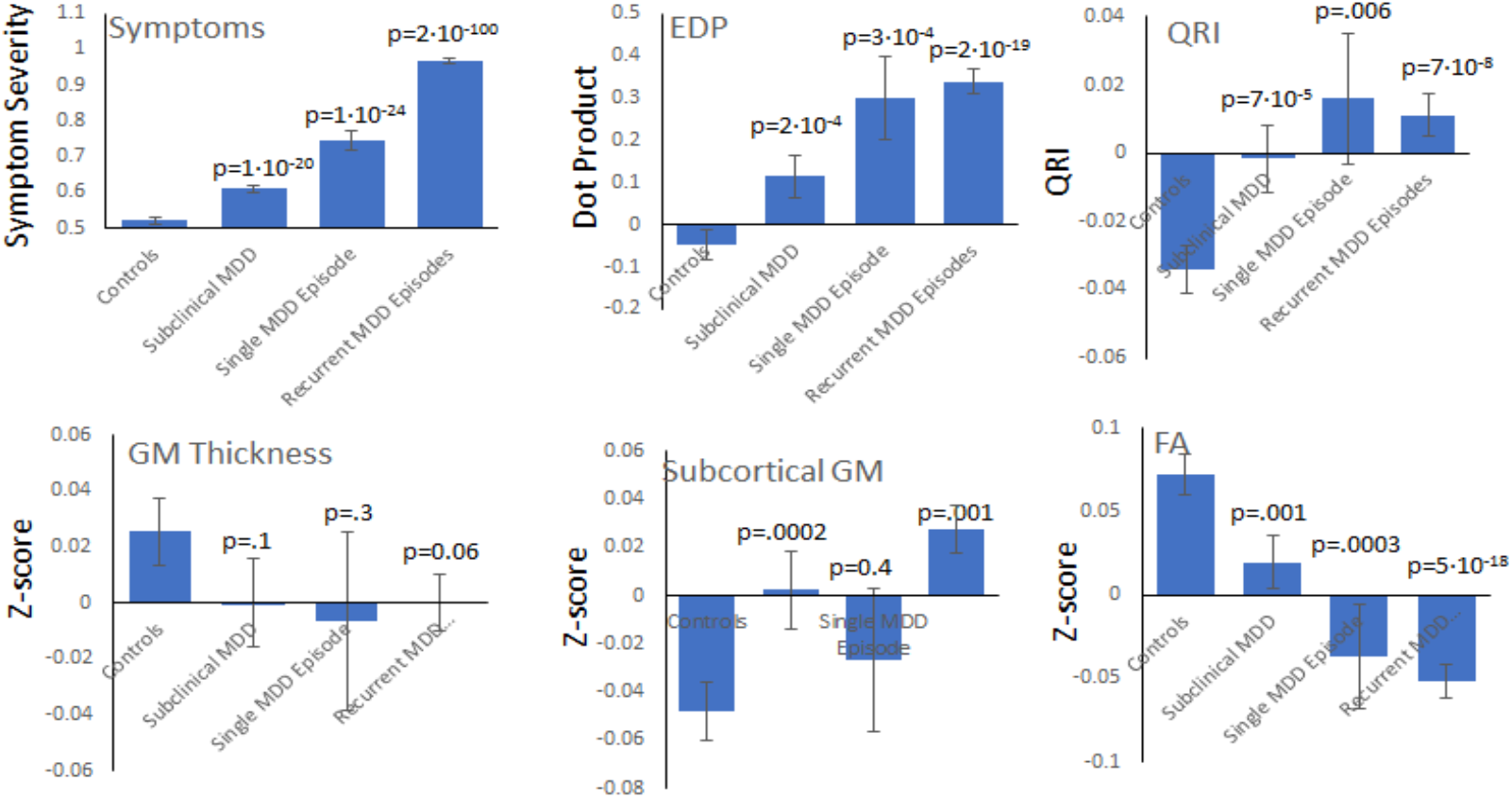
Symptom severity (0-4), EDP (normalized dot product), QRI (deviation from normal aging) and whole-brain measurements of gray matter (GM) thickness, subcortical volume and fractional anisotropy (FA) of water diffusion, expressed as normalized Z-scores after regressing age and sex effects were plotted for 4 groups – Controls (no evidence for MDD), Subclinical Depression, Single MDD Episode and Recurrent MDD groups.

### 3.2 Effects of MDD on cognition

MDD was associated with lower cognitive function consistently across all MDD groups (Figure 2). Significant differences were observed for three out of nine tasks including Matrix, reaction time (RT) and Fluid Intelligence (Figure 2) compared to healthy controls.

**Figure 2.**
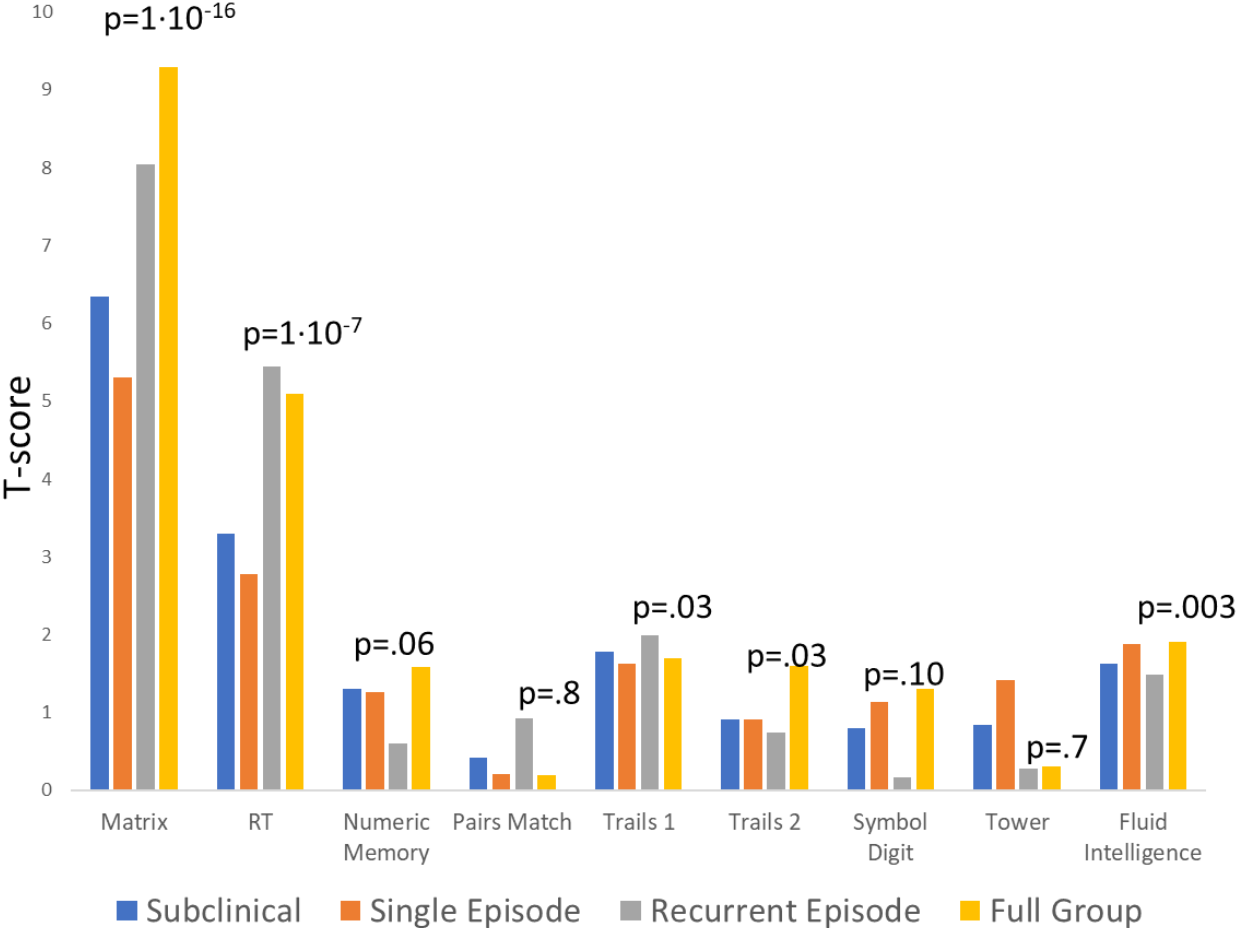
The pattern of cognitive differences (t-score) for subjects with MDD vs. controls. Significance is shown for the whole group vs. control analyses (yellow bars).

### 3.3. Cognitive association

We tested if the variance explained by a biomarker for each cognitive domain matched the pattern of clinical deficits in MDD subjects. In subjects with MDD, both EDP and GM thickness (GMT) matched (r=0.90 and 0.85, p<0.05) the pattern of association observed in MDD vs. control group differences in cognitive measures (Figure 3). However, EDP explained a numerically higher degree of variance than GMT (average t-score = 3.1±0.7 vs. 2.0±0.8, p=0.04). The other biomarkers did not show a significant correlation with the pattern of MDD-related cognitive deficits. We repeated the analysis in the controls. The t-scores of DP and GM thickness in model 1 for controls were significantly correlated with the t-scores of the MDD-control group differences (r=0.84 and 0.82, p<0.05, for DP and GM thickness, respectively) (Figure 4). Likewise, the EDP explained a higher overall proportion of the cognitive variance than GMT (average t-score = 3.6±0.7 vs. 1.9±0.8, p=0.02). There were no significant correlations for t-scores for QRI or FA or GM volume (Figure 4).

**Figure 3.**
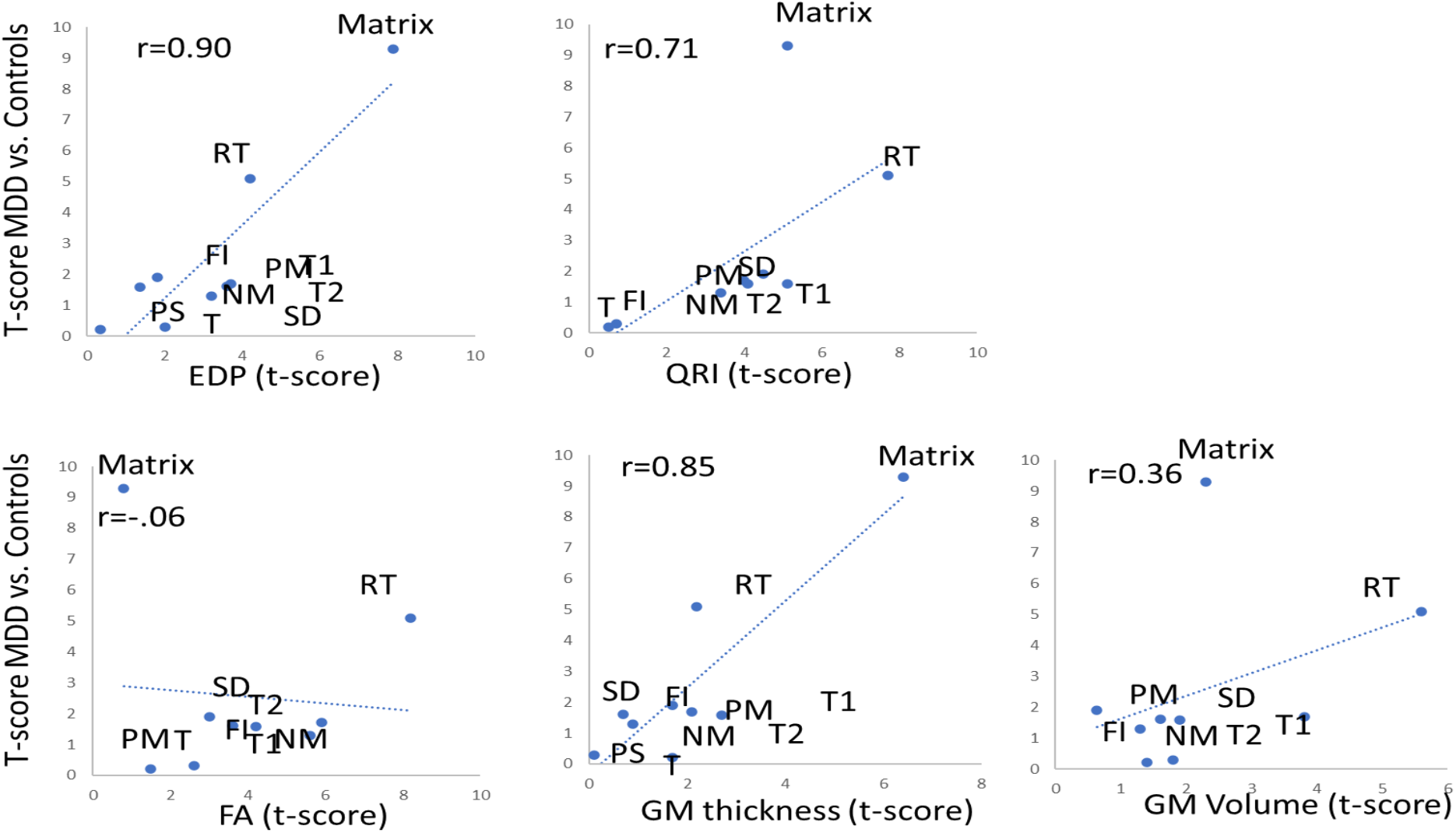
The scatter plots of the T-scores for the MDD vs. Control differences in 9 cognitive domains (Matrix, Reaction Time (RT), Numeric Memory (NM), Pairs Match (PM), Trails 1 and 2 (T1 and T2), Symbol Digit (SD), Tower (T), Fluid Intelligence (FI)) plotted versus T-scores for Model 1 predicting cognitive differences in participants affected with MDD.

**Figure 4.**
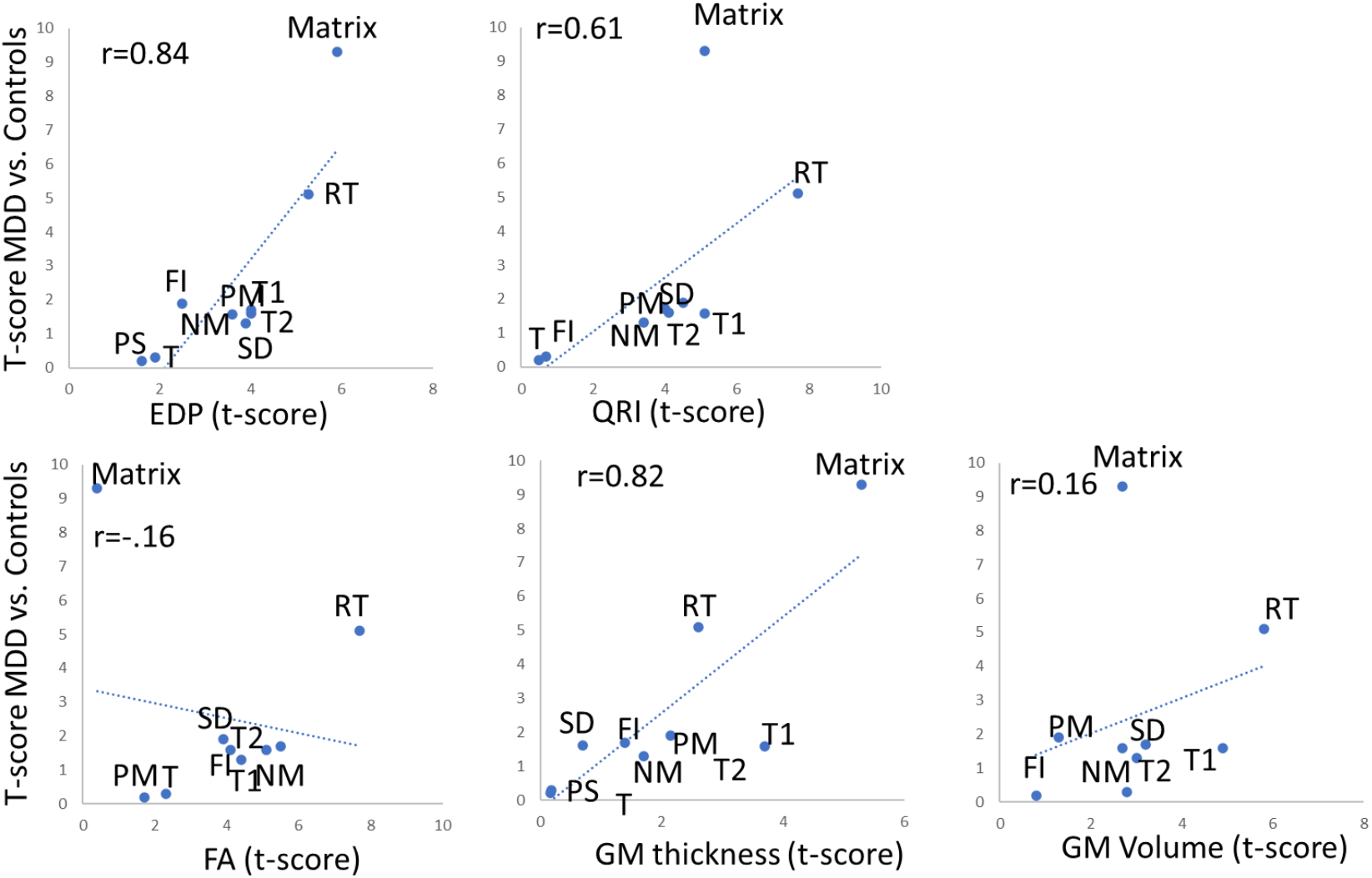
The scatter plots of T-scores for MDD vs. Control differences in 9 cognitive domains (Matrix, Reaction Time (RT), Numeric Memory (NM), Pairs Match (PM), Trails 1 and 2 (T1 and T2), Symbol Digit (SD), Tower (T), Fluid Intelligence (FI)) plotted versus T-scores for Model 1 predicting cognitive differences in normal controls.

## 4. Discussion

Using a large and inclusive dataset we evaluated the sensitivity and specificity of the ENIGMA Dop Product (EDP) index as a measurement of individual brain vulnerability for Major Depressive Disorder (MDD). EDP was calculated as a dot product between the vector of ENIGMA-MDD meta-analytic deficit values and an individual’s phenotypic vector of z scores. Higher EDP scores indicated a combination of more deviations from the expected mean and better alignment with the ENIGMA-MDD pattern. We evaluated the sensitivity and specificity of EDP to disorder severity and cognitive deficits and compared it to an index of accelerated aging (QRI) as well as whole-brain average values for gray matter (GM) thickness and volumes and fractional anisotropy (FA) of water diffusion. The EDP was superior to QRI and whole-brain measurements in separating participants with different states of MDD and healthy controls and showed the strongest association with symptoms of depression in subjects with MDD. The higher EDP values were associated with poorer performance on cognitive tasks in the MDD group. The associations were stronger in cognitive tests affected by MDD. Repeating these analyses in controls demonstrated a similar trend, suggesting that EDP is also applicable to individuals free of depression and the EDP is sensitive to cognitive performance in healthy individuals. Overall, the novel vulnerability construct, EDP, shows the ability to transfer big data neuroimaging findings to the individual and can characterize vulnerability to MDD and deficits in cognitive performance, even in non-psychiatric individuals.

Big Data neuroimaging studies have markedly improved the stability of research findings in MDD (17, 18). The ENIGMA-MDD workgroup published regional deficit patterns that were replicable across independent datasets, including the UKBB (9). Here, we performed a head-to-head comparison to demonstrate the applicability of EDP in MDD research and its superiority to other measurements including measures of accelerated aging and whole-brain average neuroimaging measurements. The EDP showed a superior ability to separate subjects with MDD based on the severity of their illness compared to all other measurements that we selected. The EDP index showed a gradual increase from controls, to subjects with subclinical MDD, to MDD and recurrent MDD. Among all neuroimaging indices, only the average FA of cerebral white matter showed an intergroup pattern that matched that. However, EDP showed a more statistically significant intergroup difference than the average FA and a stronger association with symptoms in individual subjects than FA. The follow up analyses demonstrated that while the average FA was sensitive to the symptom severity, it did not track the cognitive deficits in MDD.

To further understand the roles of EDP on specificity to MDD, we used it to explain cognitive variance in subjects with MDD and normal controls. EDP tracks similarity to brain structural patterns and therefore incorporates both genetic and developmental risk factors associated with MDD and may provide the ability to characterize the potential vulnerability for cognitive impairments in both people affected with MDD and non-psychiatric controls. Cognitive deficits are not a substantial part of the diagnostic criteria for MDD but they are prominent beyond the potential ‘impaired concentration’ criterion and are debilitating features that contribute to the socioeconomic burden (19). In the UKBB sample, subjects with MDD performed significantly more poorly than controls on the Matrix, Reaction Time and Fluid Intelligence measures - consistent with the ‘mental fog’ reported in MDD patients (20). The EDP has readily replicated that pattern of association, while the pattern for other indices - except for GM thickness - did not match that of MDD. Specifically, FA and QRI selectively explained variance in the cognitive tests sensitive to information processing speed and working memory, while GM volume was not significantly associated with cognitive performance. The cortical GM thickness explained cognitive variance in a matching pattern. This was surprising because subjects with MDD did not demonstrate significant differences in whole-brain GM thickness, nor did this biomarker show an association with symptom severity (6).

We further tested the specificity of cognitive association by repeating this analysis in controls. Our hypothesis was that people in the general population that show similarity in brain metrics, as indexed by EDP, may be at higher risk for subtle cognitive deficits. We found that higher EDP values in non-neuropsychiatric subjects were significantly and inversely associated with cognitive performance in all nine cognitive measures. The EDP was especially sensitive to and explained more variance in the cognitive tests affects by MDD. The association between EDP and cognition matched the profile of cognitive deficits in MDD (r=0.84). The cortical GM thickness was likewise associated with cognitive performance in a similar pattern. Even so, the degree of variance that it explained in either subjects with MDD or controls was significantly smaller than that explained by EDP.

This study has several limitations. The diagnostic information provided in the UKBB is based on self-report and hospital records, but is not verified by independent clinical interviews, and thus is subject to some misclassification error (24). The study is also limited as it is based on the existing UKBB sample which is cross-sectional. Expansion of these large data sets into different phases of life as well as still other brain diseases would allow for improvement upon the proposed validity of these indices.

## 5. Conclusion

The ENIGMA Dot Product is a novel anatomically informed index of vulnerability that is calculated as a similarity index between an individual’s brain patterns and expected patterns of deficits in cortical, subcortical, and white matter reported by the ENIGMA consortium. We found that higher EDP were associated with more severe symptoms of depression in subjects with MDD and with lower cognitive performance in psychiatrically healthy controls, in a pattern mirroring that observed in subjects with MDD. Indexing the similarity to brain imaging phenotypic patterns reported by big data neuroimaging studies of psychiatric conditions provides a characterization of the vulnerability to severe mental illnesses.

## Data Availability

Data is available through UKBB and software is freely available as an R package

## 6. Acknowledgement

This work was supported by the National Institutes of Health grants R01MH112180, R01MH116948, S10OD023696, R01EB015611, R01MH117601, R01AG095874, R01MH116147, and U01MH108148. These funding sources provided financial support to enable design and conduct of the study or collection, management, or analysis of the data. LS is supported by a NHMRC Career Development Fellowship (1140764) and a University of Melbourne Dame Kate Campbell fellowship. None of the funding agencies had a role in the interpretation of the data. None had a role in the preparation, review, or approval of the manuscript. None had a role in the decision to submit the manuscript for publication

